# Nutritional efficacy of *Chlorella* supplementation depends on the individual gut environment: randomized control study

**DOI:** 10.1101/2020.09.03.20184556

**Authors:** Yuichiro Nishimoto, Tatsuhiro Nomaguchi, Yuka Mori, Masaki Ito, Yuya Nakamura, Masaki Fujishima, Shinnosuke Murakami, Takuji Yamada, Shinji Fukuda

## Abstract

Recent studies have accumulated evidence that the intestinal environment is strongly correlated with host diet, which influences host health. A number of dietary products whose mechanisms of influence operate via the gut microbiota have been revealed, but they are still limited. Here, we investigated the dietary influence of *Chlorella*, a green alga commercially available as a dietary supplement. A randomized, double-blind, placebo-controlled crossover trial including 40 Japanese participants with constipation was performed and followed by integrated analysis of the gut microbiome, gut metabolome and blood parameters based on a metabologenomics approach. We revealed that the consumption of *Chlorella* increased the level of several dicarboxylic acids in faeces. Furthermore, the analysis showed that individuals with low concentrations of faecal propionate increased its concentration by *Chlorella* intake. In addition, increasing of blood folate levels were negatively correlated with defecation frequency at baseline. Our study suggested that the effect of *Chlorella* consumption varies by individuals depending on their intestinal environment, which illustrates the importance of stratified dietary management based on the intestinal environment in individuals.

## Introduction

*Chlorella* is a genus that belongs to the class Chlorophyceae and consists of mostly green algae living in freshwater. *Chlorella* species are protein-rich and currently well known as a dietary supplement that is commercially available and consumed by various populations. Recent studies have reported that *Chlorella* has various beneficial effects, such as anti-inflammatory (1) and anti-allergic activities (2), lipid metabolism improvement (3), and prevention of arteriosclerosis and cardiovascular diseases (4). *Chlorella* species also include dietary fibre. It has been reported that dietary fibre has effects such as lowering blood glucose levels, lowering cholesterol levels and suppressing intestinal inflammation (5–7). However, dietary fibre has a variety of sugar compositions, and some of its positive effects on human health depend on this composition (8). It is important to investigate the effect of each food that contains dietary fibre. In addition, the effect of dietary fibre sometimes depends on the host gut microbiota, as the gut microbiota is involved in its mechanism of action. For instance, improvement in glucose metabolism by barley consumption is associated with increased abundances of the intestinal bacterial genus *Prevotella* (5). The improvement of glucose metabolism by succinate, which is produced from dietary fibre by *Prevotella*, has been proposed as the mechanism of action (9). Rat studies have reported that *Chlorella* intake improved blood lipid profiles which correlated with gut microbiota alteration (3, 10). However, the gut microbiota is known to be different in rodents and humans (11); thus, the effect of *Chlorella* intake into the intestinal environment of humans could differ from that of rats. A previous study reported that there are inter-individual differences in the response to *Chlorella* intake (12); however, while the study focused on genetic differences among consumers, individual differences in the intestinal environment were not considered. The intestinal environment differs between human individuals (13). In this interventional study, we reculted partially constipated participants to evaluate their responses to *Chlorella* intake. In particular, this study focused on their blood parameter and intestinal environment analyzed by a metabologenomics approach combined with 16S rRNA gene-based microbiome and mass spectrometry-based metabolome analysis.

## Results

### *Chlorella* intake affected the abundances of dicarboxylic acid metabolites in faeces

To assess the effects of *Chlorella* intake, a randomized, controlled crossover trial with two 4-week dietary intervention periods separated by a 4-week washout period was performed (Fig. 1). A total of 40 Japanese participants with constipation tendencies passed the inclusion criteria, and all of the participants completed the two dietary intervention periods. Participants in two randomized blocks had similar clinical characteristics regarding the primary and secondary outcomes (defecation frequency and blood folate level) before the dietary intervention (Table S1).

**Fig. 1.**
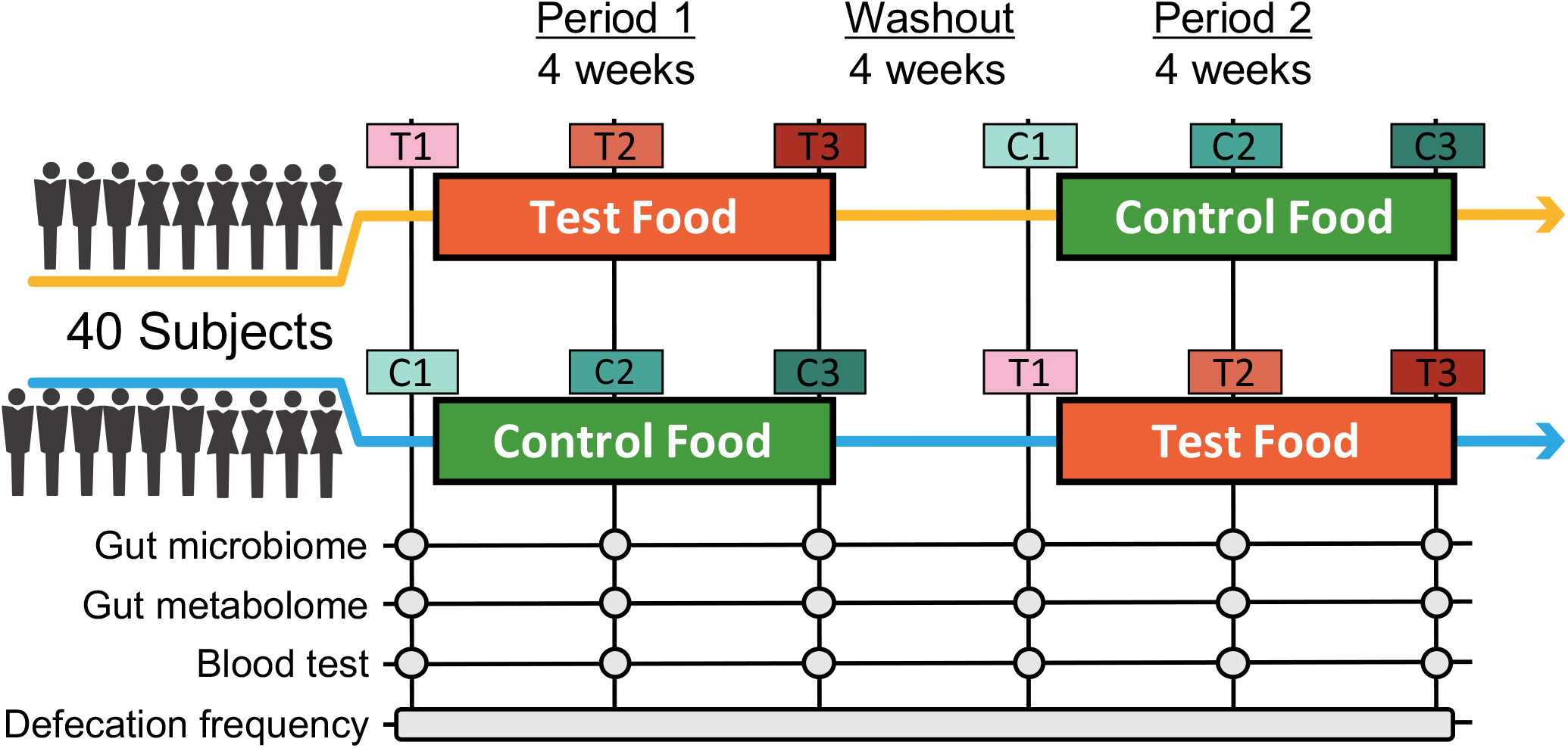
Timeline of the double-blind randomized crossover trial. Two four-week dietary treatments were set in succession. The dietary intervention periods were interspaced by a four-week washout period. Blood and stool samples were collected beforehand and at week 2 and week 4 of each intervention period. C and T indicate the control food and test food intervention periods, respectively, and 1 indicates the first sampling time point in the period.

For the primary and secondary outcomes, changes in defecation frequency and blood folate level, which are hypothesized to both be improved by *Chlorella* intake (12, 14), were analysed. Fasting blood samples were taken before and during each intervention period, and folate levels were biochemically analysed. For defecation frequency, participants were asked to record their defecation frequency during intervention periods. The defecation frequencies during days 14 - 28 in the control intervention period and the *Chlorella* intervention period were compared; however, no significant differences were observed in both defecation frequency and blood folate levels (Table 1).

**Table 1:** *Chlorella* intake had not significantly effect on primary and secondary outcomes.

|  | Effect size<sup>a</sup> | p-value<sup>b</sup> | C1<sup>c</sup> | T1<sup>c</sup> | C3<sup>c</sup> | T3<sup>c</sup> |
| --- | --- | --- | --- | --- | --- | --- |
| Defecation frequency<sup>d</sup>(/week) | 0.101<br>(-0.426 to 0.628) | 0.702 | 3.95 ± 1.05 | 4.15 ± 1.42 | 4.98 ± 1.36 | 5.30 ± 1.99 |
| Blood folate level (ng/ml) | 0.208<br>(-0.604 to 1.020) | 0.607 | 9.21 ± 4.29 | 8.48 ± 3.39 | 8.10 ± 3.76 | 7.84 ± 2.95 |

<sup>a</sup> Values are given as the mean and 95% confidence interval.
<sup>b</sup> Values indicate Welch's t-test result.
<sup>c</sup> Values are given as Mean±Standard Deviation (S.D.)
<sup>d</sup> Values represent mean of defecation frequency per week which were recorded for 2 weeks; C1 and T1 were recorded before placebo and test intervention period start, respectively. C3 and T3 were recorded during 3rd and 4th week of each intervention period.

We next performed 16S rRNA-based microbiome analysis and capillary electrophoresis time-of-flight mass spectrometry (CE-TOFMS)-based metabolome analysis. In this study, 20 randomly selected subject samples were analysed. Faecal samples were obtained prior to the intervention (C1 and T1 in Fig. 1. C and T indicate the control food and test food intervention periods, respectively, and 1 indicates the first sampling time point during the intervention) and at approximately 14 days (C2 and T2) and 28 days (C3 and T3) into each intervention period (Fig. 1). Multivariate analysis based on the Spearman rank correlation coefficient showed that the inter-individual distance in microbiome and metabolome profiles was significantly higher than the intra-individual distance (Fig. 2a, c; *p* = 3.28*10^-76^ and 4.07*10^-56^ in using microbiome and metabolome profile, respectively; Wilcoxon rank sum test). This indicated that inter-individual variation in microbiome and metabolome profiles is more prominent than the effect caused by the intervention. Therefore, to evaluate the effect of *Chlorella* intake on the microbiome and metabolome profile, it is necessary to compare variations within the individual. We compared intra-individual variation before and after placebo intake (C1 and C2, and C1 and C3) and intra-individual variation before and after *Chlorella* intake (T1 and T2, and T1 and T3), but no significant difference was detected (Fig. 2b, d). These results indicated that *Chlorella* intake had no significant effect on the general profile of the microbiome and the metabolome.

**Fig. 2.**
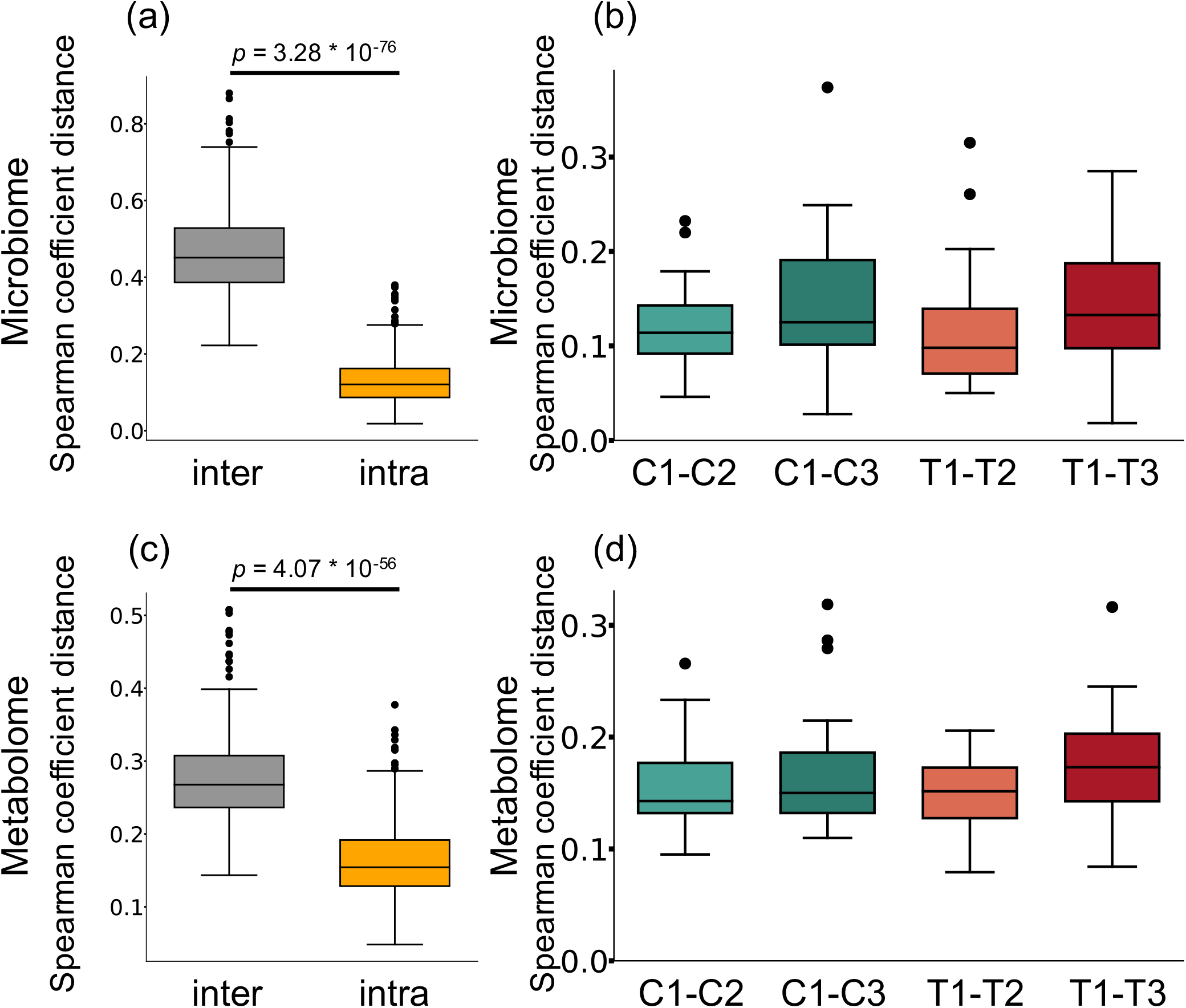
The influence of *Chlorella* consumption on the gut microbiome and metabolome profiles was not larger than that of control food consumption. (a, c): Box plot representing distribution of Spearman coefficient distance for gut microbiome (a) and metabolome profiles (c) among samples from different subjects at first time point (inter) and the distance between samples from the same subject (intra). (b, d): The boxplots show the microbiome profile distance (b) and metabolome profile distance (d) within an individual between the labelled time points. A significant difference test was performed during the same intake period (T1-T2 and C1-C2, or T1-T3 and C1-C3), but no significant difference was detected (Wilcoxon signed-rank test).

While *Chlorella* intake had no significant effect on the entire microbiome and metabolome profile, some of the specific intestinal bacteria or metabolites may have been affected. To assess the effect of intervention on each bacterium and metabolite, the relative abundance of each bacterium and scaled peak areas of each metabolite, which represent its amount in faeces, were analysed. During the analysis, we performed the Wilcoxon signed-rank test with two sets of pairs: comparison between T1 and T3 for comparison before and after *Chlorella* intake and comparison between T1-T3 and C1-C3 for comparison of the effect caused by control food and *Chlorella* intake (T1-T3 and C1-C3 represent the difference in values between the 3rd and 1st time points of the test food and the control food intervention period, respectively, in each subject). Although significant differences in bacterial abundance were detected in either one comparison or the other, there were 10 metabolites that showed significant differences in both comparisons (Fig. 3). The functions of these metabolites were analysed using the KEGG BRITE database, and the results revealed that half of these metabolites were dicarboxylic acids (Table 2). Three metabolites, azelaic acid, suberic acid, and 6-hydroxyhexanoic acid, remained significant after false discovery rate (FDR) adjustment by the Benjamini-Hochberg (BH) approach. Azelaic acid remained significant after FDR adjustment in comparison between T1 and T3; suberic acid and 6-hydroxyhexanoic acid remained significant after FDR adjustment in comparison between T1-T3 and C1-C3. *Chlorella* itself may contain these three metabolites (azelaic acid, suberic acid, 6-hydroxyhexanoic acid). Therefore, CE-TOFMS metabolic profiling was performed to measure the nutritional contents in *Chlorella*. However, these three metabolites were not contained in *Chlorella* (Table S2).

**Fig. 3.**
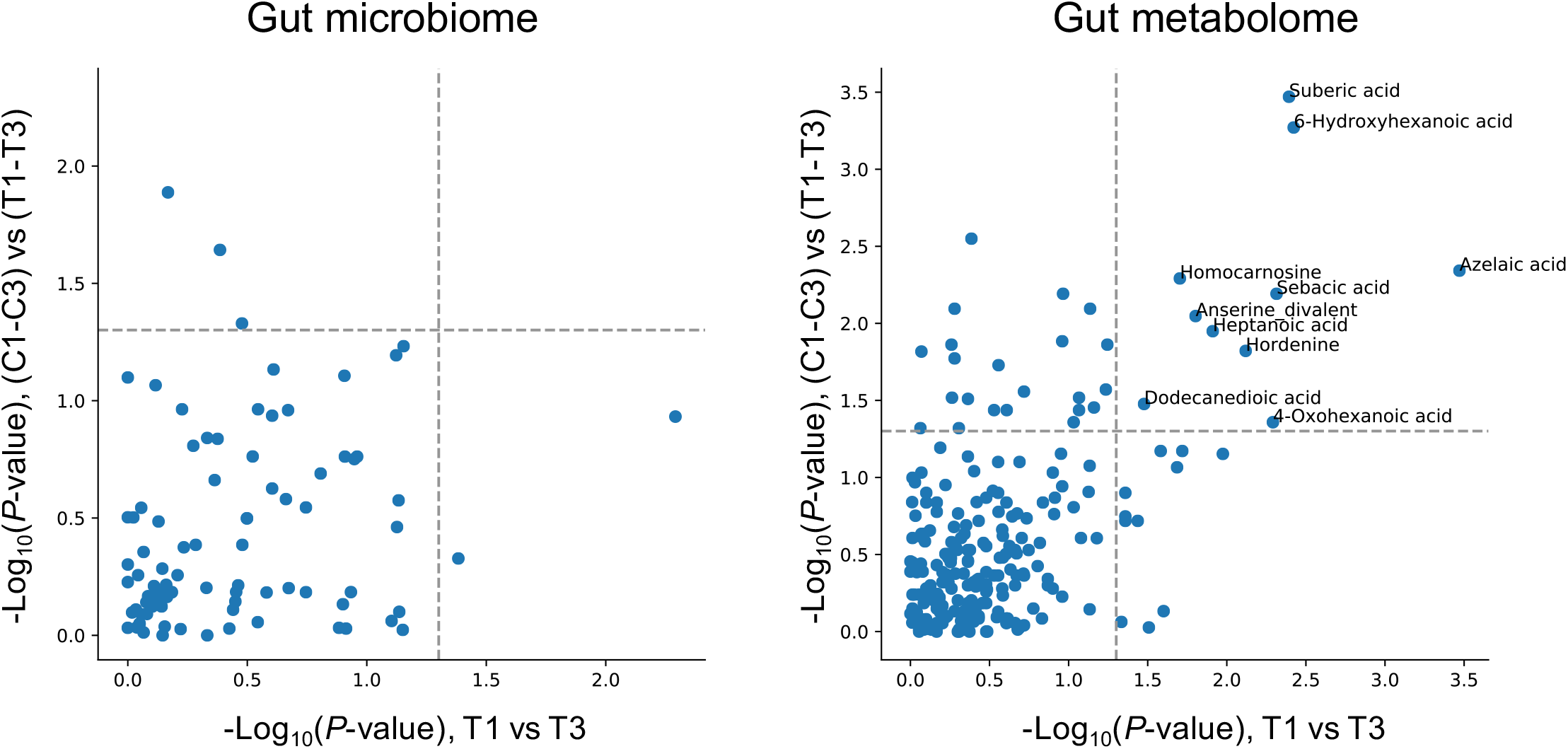
*Chlorella* intake significantly alters some intestinal metabolite levels. The X axis indicates the logarithmic *P*-value of the Wilcoxon signed-rank test for each bacterial taxon/metabolite between T1 and T3. The Y axis indicates the logarithmic *P*-value of the Wilcoxon signed-rank test for each bacterial taxon/metabolite between T1-T3 and C1-C3 (T1-T3 means differences between T1 and T3 with each subject treated as one group). Bacterial/metabolite names are labeled if a significant difference was detected in both statistical tests.

**Table 2:**
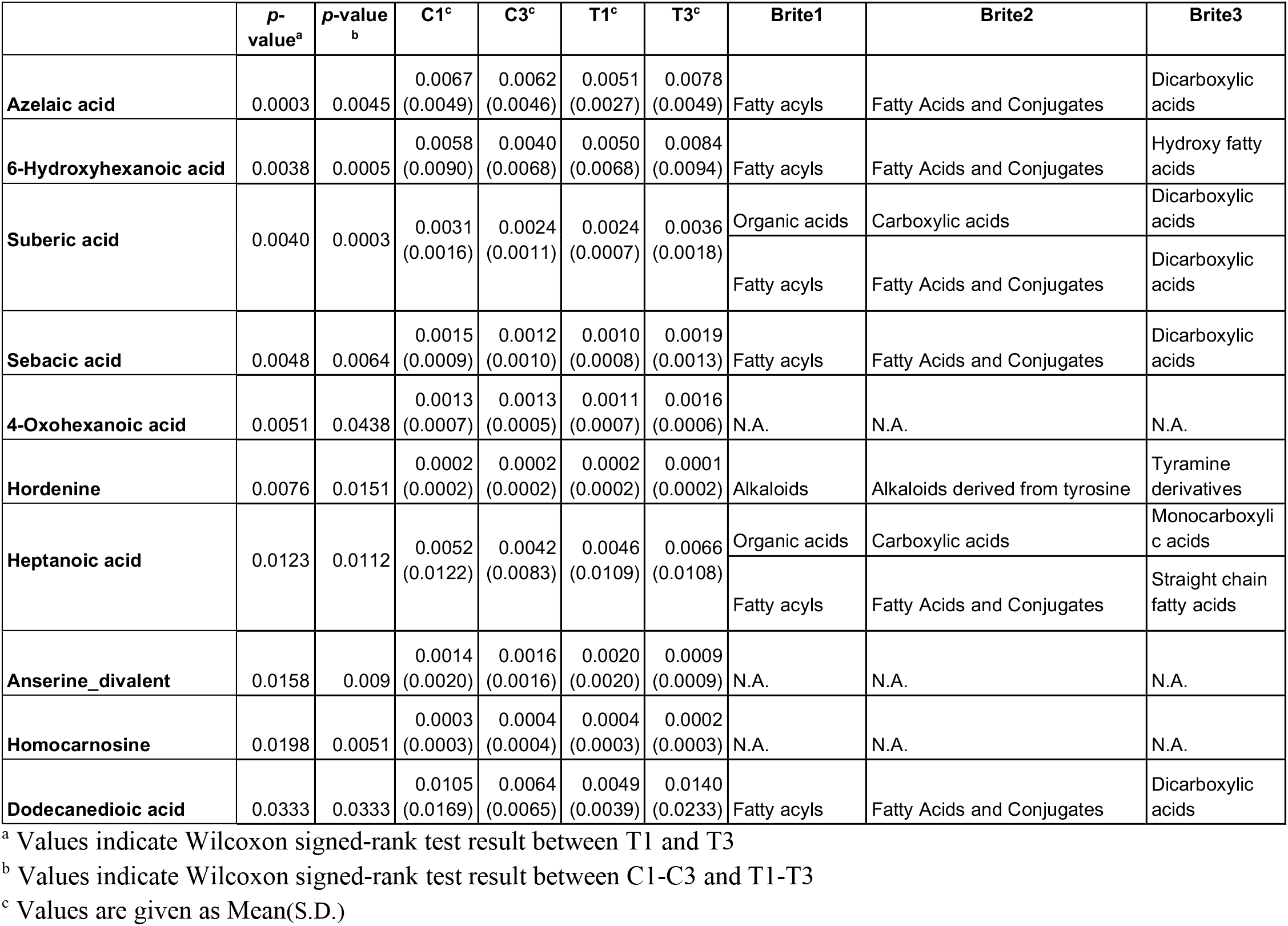
*Chlorella* intake increases dicarboxylic acid levels in faeces. Classification of significantly increased faecal metabolites upon *Chlorella* intake based on the KEGG BRITE database *(p* < 0.05 in both comparisons between C1-C3 and T1-T3 and between T1 and T3)

|  | p-value<sup>a</sup> | p-value<sup>b</sup> | C1<sup>c</sup> | C3<sup>c</sup> | T1<sup>c</sup> | T3<sup>c</sup> | Brite1 | Brite2 | Brite3 |
| --- | --- | --- | --- | --- | --- | --- | --- | --- | --- |
| **Azelaic acid** | 0.0003 | 0.0045 | 0.0067<br>(0.0049) | 0.0062<br>(0.0046) | 0.0051<br>(0.0027) | 0.0078<br>(0.0049) | Fatty acyls | Fatty Acids and Conjugates | Dicarboxylic acids |
| **6-Hydroxyhexanoic acid** | 0.0038 | 0.0005 | 0.0058<br>(0.0090) | 0.0040<br>(0.0068) | 0.0050<br>(0.0068) | 0.0084<br>(0.0094) | Fatty acyls | Fatty Acids and Conjugates | Hydroxy fatty acids |
| **Suberic acid** | 0.0040 | 0.0003 | 0.0031<br>(0.0016) | 0.0024<br>(0.0011) | 0.0024<br>(0.0007) | 0.0036<br>(0.0018) | Organic acids | Carboxylic acids | Dicarboxylic acids |
|  |  |  |  |  |  |  | Fatty acyls | Fatty Acids and Conjugates | Dicarboxylic acids |
| **Sebacic acid** | 0.0048 | 0.0064 | 0.0015<br>(0.0009) | 0.0012<br>(0.0010) | 0.0010<br>(0.0008) | 0.0019<br>(0.0013) | Fatty acyls | Fatty Acids and Conjugates | Dicarboxylic acids |
| **4-Oxohexanoic acid** | 0.0051 | 0.0438 | 0.0013<br>(0.0007) | 0.0013<br>(0.0005) | 0.0011<br>(0.0007) | 0.0016<br>(0.0006) | N.A. | N.A. | N.A. |
| **Hordenine** | 0.0076 | 0.0151 | 0.0002<br>(0.0002) | 0.0002<br>(0.0002) | 0.0002<br>(0.0002) | 0.0001<br>(0.0002) | Alkaloids | Alkaloids derived from tyrosine | Tyramine derivatives |
| **Heptanoic acid** | 0.0123 | 0.0112 | 0.0052<br>(0.0122) | 0.0042<br>(0.0083) | 0.0046<br>(0.0109) | 0.0066<br>(0.0108) | Organic acids | Carboxylic acids | Monocarboxylic acids |
|  |  |  |  |  |  |  | Fatty acyls | Fatty Acids and Conjugates | Straight chain fatty acids |
| **Anserine_divalent** | 0.0158 | 0.009 | 0.0014<br>(0.0020) | 0.0016<br>(0.0016) | 0.0020<br>(0.0020) | 0.0009<br>(0.0009) | N.A. | N.A. | N.A. |
| **Homocarnosine** | 0.0198 | 0.0051 | 0.0003<br>(0.0003) | 0.0004<br>(0.0004) | 0.0004<br>(0.0003) | 0.0002<br>(0.0003) | N.A. | N.A. | N.A. |
| **Dodecanedioic acid** | 0.0333 | 0.0333 | 0.0105<br>(0.0169) | 0.0064<br>(0.0065) | 0.0049<br>(0.0039) | 0.0140<br>(0.0233) | Fatty acyls | Fatty Acids and Conjugates | Dicarboxylic acids |

<sup>a</sup> Values indicate Wilcoxon signed-rank test result between T1 and T3
<sup>b</sup> Values indicate Wilcoxon signed-rank test result between C1-C3 and T1-T3
<sup>c</sup> Values are given as Mean(S.D.)

### Correlation analysis revealed that the effect of *Chlorella* intake depends on the individual intestinal environment

*Chlorella* contains dietary fibre that is metabolized into short-chain fatty acids (SCFAs) such as propionate and butyrate, which are known to have beneficial effects on host health, by the gut microbiota. However, a simple comparison between before and after *Chlorella* intake in this study showed no significant difference in the faecal abundances of propionate and butyrate (Fig. 4a, c). A previous study reported that the effects of probiotics on SCFAs in faeces differ between individuals depending on their SCFA levels at baseline (15). We analysed the correlation between the effect size of *Chlorella* intake, which will be referred to as the responder score (see Materials and methods for detail), and the baseline stool propionate and butyrate levels of each individual. The propionate responder score showed a negative correlation with the propionate levels at baseline (Fig. 4b; Spearman = −0.589; p = 0.00623). This indicates that subjects with a low propionate levels in faeces before *Chlorella* consumption increase in propionate after *Chlorella* intake. Butyrate also had a similar tendency, but the correlation was not significant (Fig. 4e; Spearman r = −0.362; p = 0.116).

**Fig. 4.**
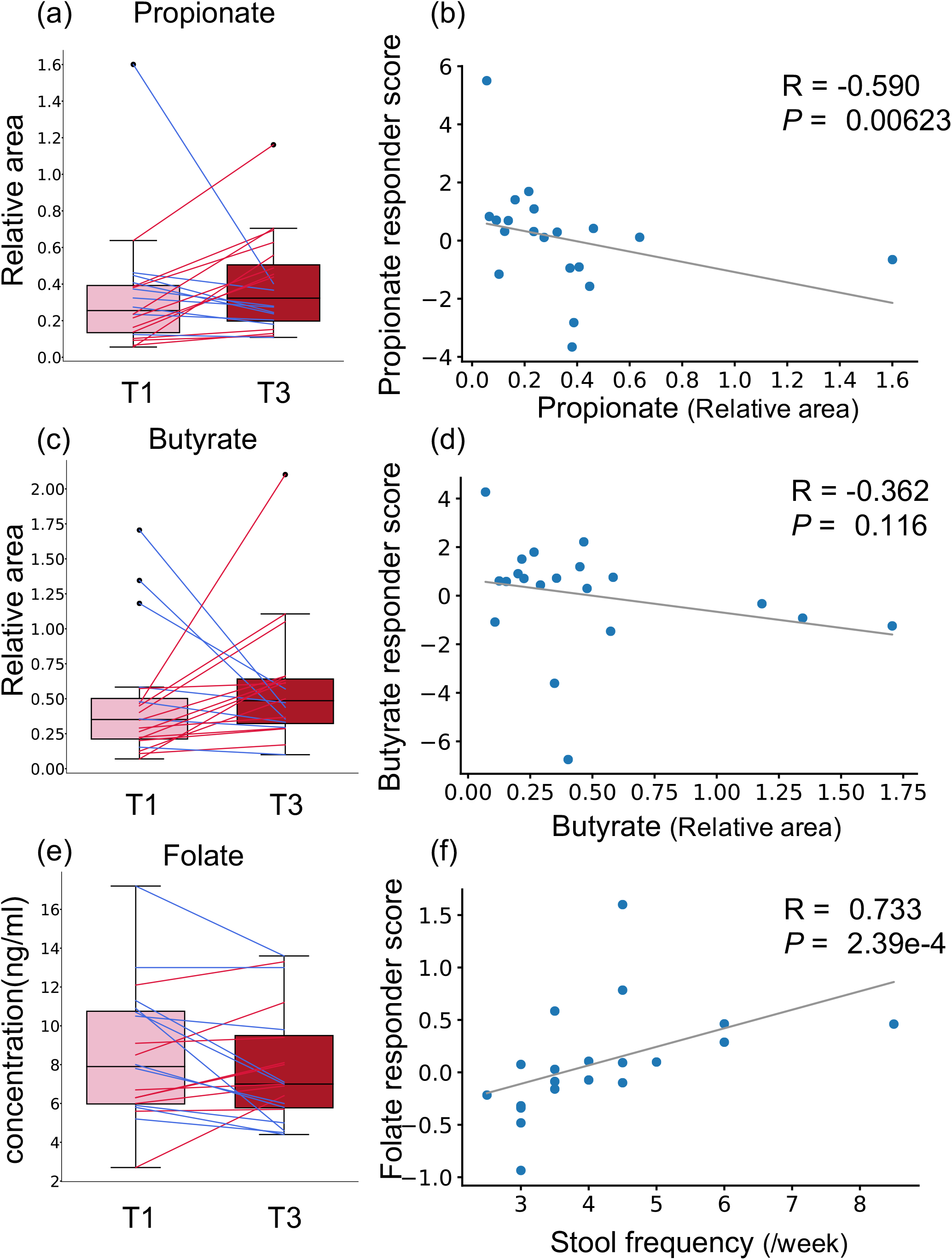
Effect of *Chlorella* intake depends on intestinal environmental factors at baseline. (a, c, e): Box plots representing distribution of faecal propionate (a), butyrate (c), and blood folate (e) concentration. No significant difference was detected (Wilcoxon signed-rank test). (b, d, f): The X axis indicates the value at baseline. The Y axis indicates the responder score of each metabolite/blood test item. Correlation was determined using the Spearman coefficient. Gray line shows the approximated curve.

SCFAs are major faecal metabolites that are produced via the metabolism of dietary fibre by the gut microbiota. Therefore, to evaluate the bacteria contributing to the response, a correlation analysis was performed on the propionate responder scores and butyrate responder scores and the abundance of each intestinal bacterial taxon at baseline. The abundance of the two bacteria *(Ruminiclostridium* 9 and *Butyricimonas)* were significantly correlated with the propionate and butyrate responder scores (Table S3). Bacteria that correlated significantly with propionate or butyrate responder score are listed in Table S3.

Next, we performed the same analysis with primary and secondary outcomes (defecation frequency and blood folate level) to evaluate whether the effect on these outcomes was also dependent on prior individual intestinal environments. To evaluate whether the effects on defecation frequency and blood folate level were dependent on the intestinal environment, correlation analysis was performed for defecation frequency responder score, blood folate responder score, and intestinal environment factors, including intestinal bacteria and metabolites abundance and defecation frequency at baseline. A strong positive correlation was found between blood folate responder score and defecation frequency at baseline (Fig. 4e, f; Spearman r = 0.733; *p* = 2.39*10^-4^). Other intestinal environmental factors that were significantly correlated with defecation frequency or blood folate responder score are listed in Table S4.

## Discussion

In our study, no significant differences in defecation frequency and blood folate level were observed between time points C3 and T3. One possible reason was the placebo effect caused by the intervention. When we compared the defecation frequency between T1 and T3 with a statistical pairwise test, a significant difference was detected. However, there was also a significant difference observed between C1 and C3, which correspond to times before and after control food intake, respectively. This suggests that placebo effects appeared during both intake periods; thus, the effect caused by *Chlorella* intake became difficult to detect when C1-C3 and T1-T3 were compared.

Multivariate analysis showed that *Chlorella* intake had no significant effect on the general profiles of either the gut microbiome or metabolome. This is the same tendency as found in our previous research (16). While the general microbiome and metabolome profiles were not significantly affected, there were several metabolites that showed significant changes in abundance after *Chlorella* intervention. Azelaic acid showed an especially clear effect since it was detected as a significantly increased metabolite even after FDR adjustment (T1-T3 comparison, q < 0.10). Azelaic acid is a commonly contained nutrient in grain such as barley. Azelaic acid can improve glucose tolerance in a mouse model (17). This effect is consistent with the result in a previous barley study that barley can also improve glucose tolerance (5). Previous studies with rodents also reported that *Chlorella* intake improves glucose tolerance (18, 19), which may be derived from an increase in intestinal azelaic acid. In this study, fasting blood glucose level did not improve significantly. Tests showing glucose tolerance, such as oral glucose tolerance tests, are needed to clarify the effect of *Chlorella* consumption.

Dicarboxylic acids, which were significantly increased metabolites in this study, were not reported to be contained in *Chlorella*. Indeed, metabolome analysis revealed that azelaic acid and suberic acid were not detected in *Chlorella* (Table S2). Therefore, the observed increase was likely derived from the metabolism of other molecules originally in *Chlorella. Chlorella* and many algae are known to produce and accumulate various long-chain fatty acid molecules (20). Such long-chain fatty acids are metabolized into dicarboxylic acids by ω-oxidation, which was also shown in an *in vivo* study in which fatty acids were administered to animals or humans that then excreted high levels of dicarboxylic acids in their urine (21). These reports suggest that the fatty acids in *Chlorella* may be metabolized into dicarboxylic acids, which increase in the blood. Dicarboxylic acids have been reported to have beneficial effects, such as antiketogenic effects; therefore, the increase in dicarboxylic acids by *Chlorella* intake would provide additional benefits other than the improvement of glucose tolerance discussed previously (22).

The effect derived from food consumption varies among individuals, and recent studies have pointed out that the intestinal environment is an important factor in determining individual dependency. For instance, barley-induced improvement in glucose metabolism is associated with an increase in the gut microbial *Prevotella/Bacteroides* ratio (5). Therefore, it is important to stratify the population into groups depending on their response to the intervention and to study those groups separately to understand the truly important elements responsible for the effect. In this study, we performed a correlation analysis with our defined responder scores and discovered that subjects with a low concentration of SCFAs increased in their faecal SCFAs, especially propionate. Although there were differences in whether the test food was probiotic or prebiotic, previous studies have shown the same tendency with the results of *Chlorella* intake (15). Propionate has beneficial effects, such as anti-inflammatory activity (23) and body weight maintenance (24). Since our data suggested that individuals with low concentrations of propionate increased its concentration by *Chlorella* intake, there is a possibility that *Chlorella* intake reduces the risk of inflammation and obesity in individuals with low concentrations of propionate. We also analysed features of the gut microbiota and metabolites in subjects with increased propionate (Table S3). There is a possibility that gut microbes that have a positive correlation with propionate and butyrate responder score may be related to produce these propionate and butyrate, and that those with a negative correlation are inhibiting propionate and butyrate production.

When we focused on the parameters related to blood folate levels, we discovered that the defecation frequency was positively correlated with the folate responder score. A previous study has shown that individuals with constipation tendencies have a lower absorption rate of nutrients than healthy individuals (25). This study showed that folate is also associated with constipation tendency. Our result may indicate that the absorbance of *Chlorella-derived* folate depends on defecation frequency.

The following limitations should be considered with our study. First, we analysed multivariate data, which included abundances of more than hundreds of intestinal bacterial taxa and metabolites. FDR correction is necessary in statistical hypothetical tests. However, the FDR correction was strict, as many items were comprehensively observed. Therefore, we discuss the factor detected by the multiple comparison without FDR adjustment. Since the factors detected by multiple comparisons without FDR adjustment possibly include false positives, validation with animal models or humans is crucial to confirm the findings. Second, this study was human intervention research that aimed to evaluate the effects of *Chlorella* intervention and elucidate the mechanism. To clarify the detailed mechanism, further study with animal models and molecular biological experiments are necessary.

In conclusion, we showed that the consumption of *Chlorella* increases the levels of several faecal dicarboxylic acids, such as azelate, which may improve glucose tolerance. On the other hand, the blood folate level, which has been reported to be increased by *Chlorella* consumption, was increased in only specific individuals with high defecation frequency. The data suggest that *Chlorella* intake with simultaneous intervention for improving bowel movement may enhance the effect of *Chlorella* consumption. In addition to the blood folate level, we discovered that the faecal propionate concentration is also improved by *Chlorella* consumption in individuals with low concentrations of propionate. Our results suggested that the effects derived from *Chlorella* consumption differ by individual based on their intestinal environments prior to intake. Such inter-individual differences in the effect of food indicate the importance of a stratified healthcare approach that involves estimating the optimal nutrition for each individual to maximize the effects of diet.

## Data Availability

The obtained sequence data are available from DRA010607, and the metabolome data are available in Tables S7

## Methods

### Ethics approval

The human rights of the subjects who participated in this study were protected at all times, and the study observed the Helsinki Declaration and the Ethical Guidelines on Epidemiological Research in Japan referring to standards for clinical trials of drugs. This trial was conducted with approval of the clinical trial ethics review committee of the Chiyoda Paramedical Care Clinic.

### Trial design and recruitment

In this study, a randomized double-blind placebo-controlled crossover trial in Japanese participants was performed for 3 months between December 2016 and April 2017 (Fig. 1, Fig. S1, Table S5). In total, 80 participants were recruited in this study. The participants fulfilled the following criteria: women aged between 20 and 59 years old with constipation (stool frequency of three to five times per week). Detailed inclusion/exclusion criteria are shown in Table S6. Based on the inclusion/exclusion criteria, 40 subjects were selected for the main trial. Randomization in this trial was performed using the block stratified randomization method. Subjects were stratified by age and stool frequency. Details about randomization were as previously reported (13). The study included 4-week dietary intervention periods in which subjects ingested test food (3 g of *Chlorella pyrenoidosa* (Sun Chlorella A Tablets®, Sun Chlorella Corp. Kyoto Japan) twice daily) and control food (3 g of digestible dextrin and pigment twice daily) in random order and separated by a 4-week washout period. In addition, faecal samples were collected at baseline (T1 and C1, T = test food and C = control food), 2 weeks (T2 and C2), and 4- weeks (T3 and C3) in the dietary intervention periods. The collected stool samples were frozen at −20°C until processing. Clinical blood tests were performed following a 12-h fasting at the same time point. In the clinical blood test, total protein, albumin, aspartate aminotransferase, alanine transaminase, lactate dehydrogenase, total bilirubin, alkaline phosphatase, γ-glutamyl transpeptidase, creatine phosphate enzyme, urea nitrogen, creatinine, uric acid, sodium, chlorine, potassium, calcium, total cholesterol, low-density lipoprotein cholesterol, high-density lipoprotein cholesterol, neutral fat, glucose, white blood cells, red blood cells, haemoglobin, haematocrit, platelet, folate, vitamin B12 and homocysteine were measured. All subjects completed the trial, and microbiome and metabolome analyses were performed for 20 out of 40 people for financial reasons. The primary outcome was stool frequency, and the key secondary outcomes were stool 16S rRNA metagenomic analysis, metabolome analysis and blood folate concentration.

### Faecal DNA and metabolite extraction

DNA extraction from faecal samples was performed as previously reported (26). After extraction, the V1-V2 variable region of the 16S rRNA gene was amplified using the bacterial universal primers 27F-mod (5'-AGRGTTTGATYMTGGCTCAG-3) and 338R (5-TGCTGCCTCCCGTAGGAGT-3) with Tks Gflex DNA polymerase (TaKaRa Bio Inc., Japan) (27). The amplicon DNA was sequenced using MiSeq (Illumina, USA) according to the manufacturer’s protocol. Extraction of metabolites from faecal samples was performed as previously reported (28). The obtained sequence data are available from DRA010607, and the metabolome data are available in Table S7.

### *Chlorella* metabolomics analysis

CE-TOFMS metabolic profiling was performed to measure the nutritional contents in *Chlorella*. 50.4 mg of powdered *Chlorella* was vortex mixed with Mili-Q water for one minute and then agitated at 500 rpm, 37°C for one hour. Samples were then centrifuged at 9,100 × *g* for 5 minutes. 250 ¼L of supernatant was ultrafiltrated using a ultrafree 5kDa MWCO centrifugal filter unit (Millipore) at 9,100 ×*g* for 80 minutes. Internal standard was added to the filtrate and analyzed using CE-TOFMS in both positive and negative mode. As a blank sample, sample without *Chlorella* powder was also prepared and analyzed.

### Bioinformatics and statistical analysis

For 16S rRNA gene analysis, QIIME2 (version 2019.10) was used (29). In the analytical pipeline, sequence data were processed by using the DADA2 pipeline for quality filtering and denoising (options: --p-trim-left-f 20 --p-trim-left-r 19 --p-trunc-len-f 240 --p-trunc-len-r 140) (30). The filtered output sequences were assigned to taxa by using the "qiime feature-classifier classify-sklearn" command with the default parameters. Silva SSU Ref Nr 99 (version 132) was used as a reference database for taxonomy assignment. In statistical analysis, the paired t-test and paired Cohen’s d in the R package effsize were used for pairwise comparison in primary and secondary outcomes. Other statistical analyses were performed with in-house Python scripts (version 3.7.3). For pairwise comparison of the relative abundances of intestinal bacterial taxa and the relative areas of intestinal metabolites, the Wilcoxon signed-rank test with Benjamini-Hochberg false discovery rate (FDR-BH) correction was used (scipy version 1.3.1 and statsmodels 0.10.1 were used for the Wilcoxon signed-rank test and FDR-BH correction, respectively). During the comparison, bacterial taxa with a mean relative abundance below 0.001 and metabolite not detected in 75% of samples were excluded.

### Defining the responder score with a specific response

In this study, the test food effect size was defined as the responder score and used to evaluate whether effects depended on individual basal characteristics. The response score was calculated with the following equation:

*Responder Score* = ((*T*3 − *T*1) *−* (*C*3 *− C*1)) */ Average*(*C*1*, T*1)

